# Mortality Outcomes in Task-Sharing for Emergency Care: Impact of Emergency Physician Supervision on Non-Physician Emergency Care in Rural Uganda

**DOI:** 10.1101/2021.09.15.21263465

**Authors:** Brian Rice, Ashley Pickering, Colleen Laurence, Prisca Mary Kizito, Rebecca Leff, Steven Jonathan Kisingiri, Charles Ndyamwijuka, Serena Nakato, Lema Felix Adriko, Mark Bisanzo

**Author notes:** Corresponding Author: Ashley Pickering, MD, MPH, University of Maryland Medical Center, Department of Emergency Medicine, 110 S. Paca St, Suite 600, Baltimore, MD 21201. **Contributorship statement:** BR, AP and MB planned the study. BR, CL analyzed the data. BR, AP, CL, PK, RL, SK, CN, SN, LF, MB interpreted the data in the local context. BR, AP, CL, RL drafted the manuscript. BR, AP, CL, PK, RL, SK, CN, SN, LF, MB revised the manuscript. AP submitted the study. BR and AP are the guarantors of the overall content. **Competing Interests:** None of the authors have competing interest to disclose. **Funding:** This study was not funded.

## Abstract

**Introduction:** Emergency care (EC) capacity is limited by physician shortages in low- and middle-income countries like Uganda. Task-sharing — delegating tasks to more narrowly trained cadres — including EC nonphysician clinicians (NPCs) is a proposed solution. However, little data exists to guide emergency medicine (EM) physician supervision of NPCs. This study’s objective was to assess the mortality impact of decreasing EM physician supervision of EC NPCs.

**Methods:** Retrospective analysis of prospectively collected data from an EC NPC training program in rural Uganda included three cohorts: “Direct” (2009-2010): EM physicians supervised all NPC care; “Indirect” (2010-2015): NPCs consulted EM physicians on an ad hoc basis; “Independent” (2015-2019): NPC care without EM physician supervision. Multivariable logistic regression analysis of three-day mortality included demographics, vital signs, co-morbidities and supervision. Sensitivity analysis stratified patients by numbers of abnormal vital signs.

**Results:** Overall, 38,344 ED visits met inclusion criteria. From the “Direct” to the “Unsupervised” period patients with ≥3 abnormal vitals (25.2% to 10.2%, p<0.001) and overall mortality (3.8% to 2.7%, p<0.001) decreased significantly. “Indirect” and “Independent” supervision were independently associated with increased mortality compared to “Direct” supervision (“Indirect” Odds Ratio (OR)=1.49 [95%CI 1.07 - 2.09], “Independent” OR=1.76 [95%CI 1.09 - 2.86]). The 86.2% of patients with zero, one or two abnormal vitals had similar mortality across cohorts, but the 13.8% of patients with ≥3 abnormal vitals had significantly reduced mortality with “Direct” supervision (“Indirect” OR=1.75 [95%CI 1.08 - 2.85], “Independent” (OR=2.14 [95%CI 1.05 - 4.34]).

**Conclusion:** “Direct” EM physician supervision of NPC care significantly reduced overall mortality as the highest risk ∼10% of patients had nearly 50% reduction in mortality. However, for the other ∼90% of ED visits, independent EC NPC care had similar mortality outcomes as directly supervised care, suggesting a synergistic model could address current staffing shortages limiting EC access and quality.

**SUMMARY BOX:** *What is already known?:*

- Physician shortages and lack of specialty training limit implementation of emergency care and associated reductions in mortality in low- and middle-income countries (LMIC) such as Uganda.
- Task-sharing, often to non-physician clinicians, is proposed as a solution however data to support safe, effective training and physician supervision protocols is limited.

*What are the new findings?:*

- The highest risk 10% of emergency care patients have approximately a 50% reduction in mortality when non-physician clinicians are directly supervised by emergency medicine physicians.
- For most emergency care patients (the lowest risk 90%) independent emergency care by non-physician clinicians provides similar morality outcomes to direct supervision by an emergency medicine physician.

*What do the new findings imply?:*

- Training of both emergency care physicians and non-physician clinicians is essential, as physicians provide improved mortality outcomes, especially for the critically ill, and non-physician clinicians will help address lack of trained and available emergency care providers in a timely, cost-effective manner.
- Physician supervision of all emergency care is the penultimate goal, however non-physician clinicians can be trained to provide comparable morality outcomes for the vast majority of patients when practicing independently.
- Triage protocols are needed to identify high-risk emergency care patients, such as those with 3 or more abnormal vital signs, for early involvement of an emergency physician either directly, or through supervision of a non-physician clinician.

## INTRODUCTION

Global recognition of the need to develop emergency care (EC) is growing.[1,2] In low- and middle-income countries LMICs, physician shortages make the provision of medical care and in particular EC problematic, with the greatest challenge centered in Sub-Saharan Africa.[3– 5] EC needs remain largely unmet throughout many LMICs, including Uganda.[5–8] Based on the estimate that 57% of deaths occurring in low-income countries are from conditions treatable with EC, approximately 160,000 Ugandans’ lives could have been saved by provision of EC in 2019.[9,10] EC in Uganda is largely limited by physician shortages, as there are 1.68 physicians per 10,000 people, amongst the lowest rates worldwide.[11] Uganda has placed a priority on developing EC over the next five years, estimating that 454 specialist EC physicians will be required by 2025.[12] EC specialty training in Uganda began in 2017 and currently certifies between five and 10 Ugandan emergency medicine (EM) specialists per year.[13] This leaves an enormous training gap with 45 and 90 years of training needed to produce EM specialists to meet the projected five-year staffing demands.

One solution to address this shortage that has been widely advocated and implemented in SSA is “task-sharing,” or delegating tasks to more narrowly trained cohorts of new or existing providers, often non-physician clinicians (NPCs).[14–20] The World Health Organization advocates for NPCs that are “adequately trained, supported and supervised”.[18,19] Though NPCs are currently providing surgical specialty, obstetric, and HIV care throughout SSA [21– 27], there has been limited application of NPC cadres to offset EC shortages.[20,28,29] High-income countries have compensated for regionally inadequate physician numbers and uneven distribution of emergency physicians by adopting physician supervised NPCs in larger emergency units and in some cases NPC practice with remote physician supervision in smaller rural hospitals.[30–33] Data and protocols to guide implementation of EC NPC training and practice in LMICs, where EM is largely newly developing, and EM physicians are typically not available, is highly limited.

Few articles exist addressing training of NPCs for roles in the African acute care settings outside of trainings focused on specific obstetric, surgical or anesthesia procedures [34–37], while others find that emergency and acute care training is lacking in NPC education in many SSA countries including Uganda.[34,38] While our research group has published on an EC NPC training program and its associated outcomes, we are not aware of any additional studies documenting a comprehensive EC NCP training program in a LMIC.[29,39–44] There are documentation a few short-courses designed to teach NCP EC skills in SSA.[45–48] Consistent with this limited evidence base, no standards exist describing if, when or how to transition to reduced supervision or independent NPC care following initial training. The impact of transitioning to decreased levels of supervision on quality of NPC care and patient outcomes is therefore unknown. This represents a major limitation in the ability to implement NPC training, supervision and uptake into health systems in a safe, effective and evidence-based manner.

While health systems are evolving in Uganda over the last decade so too is the health of the general population. Uganda’s national crude death rates decreased by 63% across all age groups (10.2/1000 in 2009 to 6.4/1000 persons in 2019) during the study period.[49] Likewise, under-five mortality decreased by approximately 38,000 deaths per year (112,747 in 2009 to 74,053 in 2019).[49] Concurrently, life expectancy increased by 6.8 years and rates of malaria and HIV infection decreased.[49]

EC has been delivered by NPCs in Uganda since 2009 in a training program that has transitioned from directly supervised to independent NPC care. The objective of this study is to compare the mortality outcomes of emergency unit patients receiving EC by NPCs during eras of direct EM physician supervision, indirect EM physician supervision, and independent NPC care while the baseline population health in Uganda was evolving. It is hoped these findings will contribute to larger policy conversations to define when and how collaboration between physicians and NPCs can produce the highest quality EC outcomes for the greatest number of Ugandans.

## METHODS

### Description of Study Site

All data comes from the emergency unit at Karoli Lwanga Hospital, a rural district hospital located in the town of Nyakibale in the Rukungiri District of southwest Uganda. The hospital has a six-bed emergency unit that opened in 2008 and treats 300 to 700 patients per month arriving between 8:00 am and midnight every day of the year. Since 2009, the emergency unit has been staffed by non-physician clinicians called emergency care practitioners (ECPs) who received training from emergency medicine physicians working with Global Emergency Care. The ECPs are nurses who have completed a two-year advanced training course in emergency care described in detail elsewhere by Hammerstedt et al (29). While the course is currently run in conjunction with MUST, the NPCs in this cohort study were trained as part of the pilot program that began through a collaboration between GEC and Karoli Lwanga Hospital. Global Emergency Care (GEC), a US-based 501(c) [3] non-governmental organization, has run a two-year EC specialty NPC training program since 2009, and currently does so in collaboration with Mbarara University of Science and Technology (MUST).

Supervision of the ECPs changed over time generating three cohorts designated “Direct Supervision”, “Indirect Supervision” and “Independent Care”. “Direct Supervision” occurred from November 2009 - April 2010 when a single US-trained EM physician practicing with a Ugandan license was on site every day and directly supervised ECP care and supplemented with clinical care in a model similar to US residency training. “Indirect Supervision” occurred from July 2010 - November 2015. During this era a volunteer US-trained EM physician was on site for approximately 85% of the weeks; however, they were present in a teaching role only and provided no direct patient care. They were available for consultation on an ad hoc basis by the ECPs and consultation was largely based on ECP discretion. “Independent Care” occurred from December 2015 - December 2019, and ECPs provided clinical care without any EPs onsite. During all periods, no Ugandan physicians were assigned to the emergency unit. Hospital physicians were similarly available throughout the study period for consultation for patients who required surgery, did not respond to initial treatments, or in whom there was considerable diagnostic uncertainty. Throughout the study period, ECPs admitted patients to the same hospital medical and surgical wards, which were staffed by Ugandan physicians with standard levels of training and no connection to GEC. Resource availability was constant over the study period and with resource utilization by clinicians in this emergency unit described in detail elsewhere.[39]

### Patients and Public Involvement

The ECP training program was originally developed in response to several years of clinical emergency medicine experience and ongoing healthcare staffing shortages in Uganda. The positive response of patients, staff and administrators at Karoli Lwanga Hospital to the training program and their interest in improving patient care led to ongoing research and program evaluation. Patients and the public were not involved in the design of the study; however, outcome measures are explicitly patient-oriented. Results have been and will continue to be disseminated through open access publication to allow local clinicians, administrators, policymakers and researchers to benefit

### Data Collection

GEC maintained a group of trained research associates who prospectively collected quality assurance data on all Karoli Lwanga Hospital emergency unit patient visits. Collected data included demographics, vital signs, laboratory and radiology testing, disposition, as well as three-day follow-up vital status (mortality) for all admitted and discharged patients. On the third day following initial evaluation in the emergency unit, patients admitted to the hospital were visited in person, and patients discharged from the emergency unit or ward were contacted via phone. This follow-up protocol included seven consecutive days of calling all patients on the phone (if they had a phone) before considering them lost to follow-up and is described in detail elsewhere. (29) Trained research assistants entered data using Microsoft Excel from 1 January 2010 – 23 March 2012, and Microsoft Access from 24 March 2012 – 31 December 2019.

### Research Ethics Approval

Ethics approval for the quality assurance database was obtained through the Institutional Review Board at Mbarara University of Science and Technology.

### Data Analysis

A cohort study was done using retrospective analysis of prospectively collected data abstracted from the Karoli Lwanga Hospital emergency unit quality assurance database, including all consecutive patients presenting to the emergency unit from November 2009 until December 2019. All patients missing age, gender, disposition and three-day follow up were excluded from analysis. Patients who were dead on arrival (lacked vital signs with no resuscitation or interventions attempted) and patients who were transferred or left against medical advice did not receive follow-up by protocol and thus were also excluded from analysis. All other patients of all ages and dispositions were included. Vital signs were taken for all patients, but the inconsistent availability of pediatric sphygmomanometers meant that blood pressures were missing for 89% of children under 5 and 47% of children aged 5-12, as compared to 3% missing in all other age groups. To control for this effect, all patients aged 12 or less that had missing blood pressure were coded as normal for analysis. No other missing variables were coded as normal, and no other data was imputed. Data was abstracted, cleaned, and analyzed by a single researcher (BR) using Stata 16.1 (StataCorp, College Station, TX).

Continuous variables were tested for significance using one-way ANOVA and proportions were compared using chi-squared. A multiple variable logistic regression model was developed to test the significance of associations between independent variables and mortality in emergency unit patients. Because only two months of data existed for 2009, and three months of data were missing for 2010, the years 2009 and 2010 were both coded as 2010 for the continuous “Year” variable included in that model. Area Under Receiver Operating Characteristics Curve (AUROC), Hosmer-Lemeshow Goodness of Fit, and Brier score were all calculated for this model.

## RESULTS

Overall, 49,804 patient visits occurred from 2009 - 2019, and 38,344 met criteria for inclusion for analysis. Inclusion and exclusion criteria are shown in Figure 1 below.

**Figure 1:** Patient Flow Diagram.

The patient characteristics of those visits stratified by eras of supervision (as described in Methods above) are shown in Table 1 below.

**Table 1:** Patient Characteristics.

| **Table 1: Patient Characteristics** |  |  |  |  |
| --- | --- | --- | --- | --- |
| **Characteristic** | **Direct Era**<br>**(n=2017)** | **Indirect Era**<br>**(n=21210)** | **Independent Era**<br>**(n=15111)** | **P-**<br>**value** |
| **Age, mean (SD)** | 25.8 (23.2) | 28.8 (24.1) | 32.9 (24.9) | < 0.001* |
| **Age Group** |  |  |  |  |
| Under 5, % (n) | 25.8% (521) | 21.0% (4446) | 14.5% (2193) | < 0.001 |
| 5-12 y.o., % (n) | 7.4% (149) | 7.7% (1622) | 7.3% (1103) |  |
| 12-18 y.o., % (n) | 8.2% (165) | 8.3% (1750) | 7.4% (1112) |  |
| 18-64 y.o., % (n) | 49.6% (1000) | 51.9% (11004) | 56.6% (8550) |  |
| > = 65 y.o., % (n) | 9.0% (182) | 11.3% (2394) | 14.3% (2153) |  |
| **HIV-positive (known status or newly diagnosed), % (n)** | 1.7% (35) | 5.6% (1185) | 6.9% (1047) | < 0.001 |
| **Malaria parasites on blood smear, % (n)** | 22.8% (460) | 18.4% (3898) | 5.6% (846) | < 0.001 |
| **Gender - Female, % (n)** | 46.5% (939) | 46.1% (9789) | 46.6% (7047) | 0.641 |
| **Complete Vital Signs, %(n)** | 87.2% (1758) | 87.5% (18571) | 90.3% (13652) | <0.001 |
| **Vital Sign Abnormalities** |  |  |  |  |
| Hypoxia, % (n) | 12.4% (250) | 11.9% (2524) | 12.7% (1917) | <0.001 |
| Tachypnea, % (n) | 51.3% (1035) | 43.1% (9140) | 27.7% (4185) | <0.001 |
| Bradycardia, % (n) | 1.8% (37) | 4.0% (856) | 4.5% (673) | <0.001 |
| Tachycardia, % (n) | 31.3% (632) | 18.3% (3888) | 15.2% (2303) | <0.001 |
| Hypothermia, % (n) | 35.9% (724) | 27.4% (5806) | 29.4% (4448) | <0.001 |
| Febrile, % (n) | 21.1% (426) | 15.6% (3300) | 13.6% (2052) | <0.001 |
| Hypotension, % (n) | 19.6% (396) | 11.9% (2533) | 7.9% (1191) | <0.001 |
| **Number of abnormal vital signs** |  |  |  |  |
| 0 or 1, % (n) | 46.2% (932) | 61.3% (13001) | 69.0% (10429) | <0.001 |
| 2, % (n) | 28.6% (577) | 23.4% (4968) | 20.8% (3136) |  |
| 3 or more, % (n) | 25.2% (508) | 15.3% (3247) | 10.2% (1546) |  |
| * ANOVA used for significance test, all others use chi-squared |  |  |  |  |

There were significant differences in every characteristic across the supervision eras except for gender. Fewer pediatric patients and thus more adult and elderly patients, fewer patients with malaria, more patients with HIV, more complete vital signs and fewer abnormal vital signs were present as the study progressed. The rates of missing vital sign data were: blood pressure 2.0%, respiratory rate 4.9%, pulse oximetry 5.7%, heart rate 1.6%, and temperature 2.1%.

The overall three-day mortality across the program from 2009-2019 was 3.1% (1,199 deaths overall). Overall, mortality increased significantly and monotonically based on the number of abnormal vital signs patients had on presentation from zero or one (1.31%, n=319 deaths in 24,362 patients), to two (3.97%, n=345 deaths in 8,681 patients) to three or more (10.1%, n=535 deaths in 5,301 patients). A clear trend towards lower mortality and a lower proportion of patients presenting with abnormal vital signs existed over time and is shown in Figure 2.

**Figure 2:** Mortality and vital sign abnormalities 2009 - 2019.

Crude mortality decreased significantly across supervision eras (Direct: 3.8%, Indirect: 3.4%, Independent: 2.7%, p<0.001). Conversely, the proportion of patients presenting with zero abnormal vitals increased across supervision eras and those presenting with three or more abnormal vital signs decreased.

Mortality across supervision eras was stratified by the number of abnormal vital signs and is reported as Appendix 1 and is displayed visually in Figure 3 below.

**Figure 3:** Mortality stratified by number of abnormal vital signs across supervision eras.

As illustrated in Figure 3, the mortality for the indirect and independent eras were very similar, while direct supervision appeared significantly different, with higher mortality in patients with zero or one abnormal vital sign and lower mortality in patients with three or more abnormal vital signs. Confidence intervals were wide, given the overall small number of fatalities during the direct era, but both differences (increased and decreased mortality) achieved statistical significance.

Given the changing baseline in patient mortality and vital sign abnormalities, a logistic regression model was developed to determine whether supervision of NPC care increased or decreased mortality for emergency unit patients. The results of this model are displayed in Table 2 below.

**Table 2:**
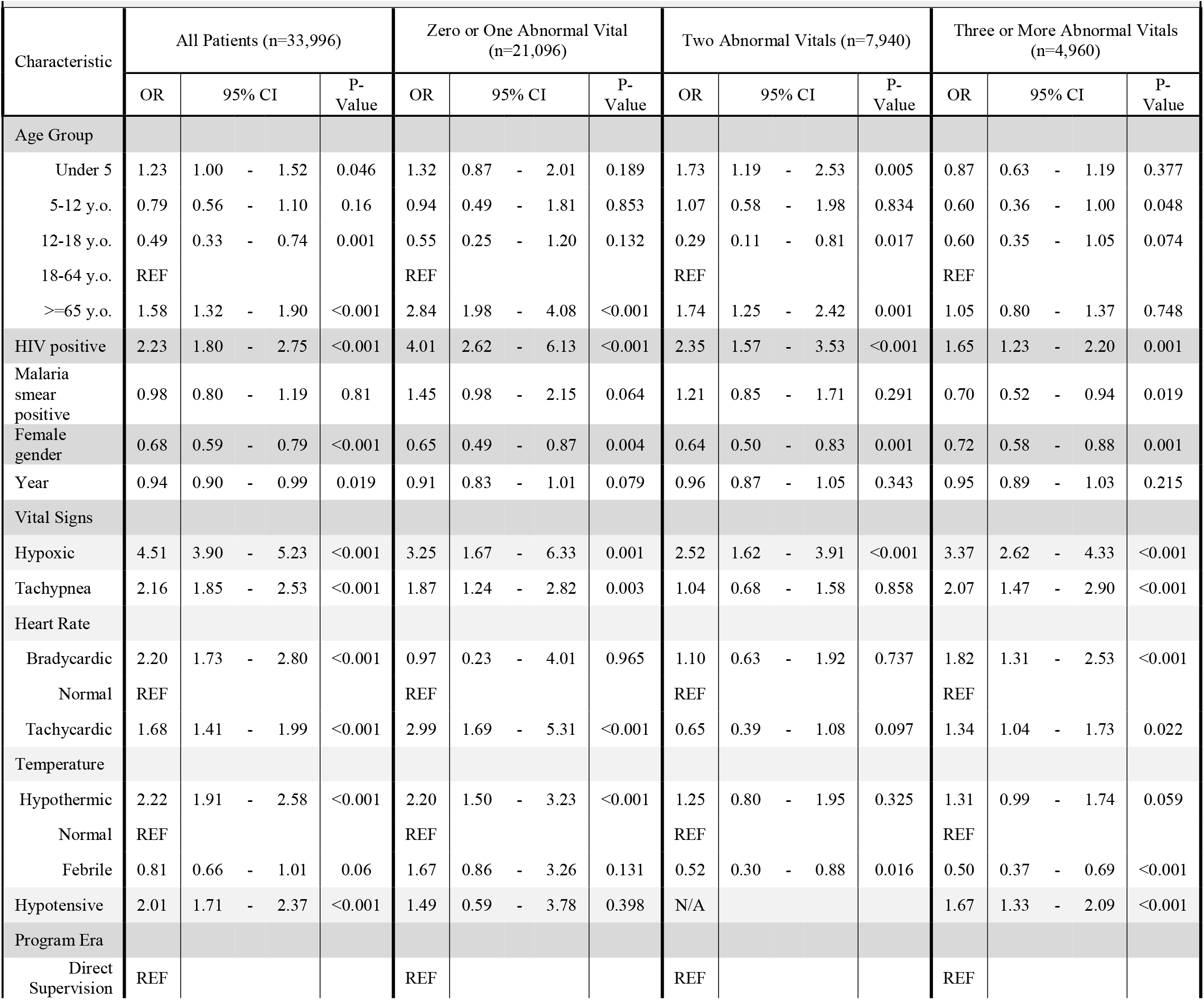
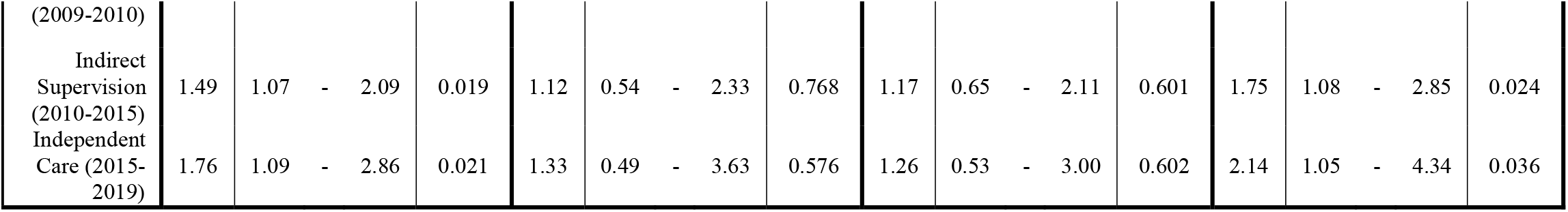
Logistic Regression

In this model, indirect supervision and independent care were both associated with increased mortality as compared to direct supervision (Indirect: Odds Ratio (OR)=1.49 [95%CI - 2.09], Independent: OR=1.76 [95%CI 1.09 - 2.86]). This model was well-calibrated (brier score 0.025), discriminated well between patients at risk for our outcome of interest (death) (AUROC 0.81) and was not over-fitted (Hosmer-Lemeshow 0.28).

As a sensitivity analysis, we looked at patients grouped by number of abnormal vital signs, as in Table 2 above. Patients with zero, one or two abnormal vital signs had no significant mortality benefit from supervision. Patients with three or more abnormal vital signs had increased mortality when comparing directly supervised NPC care to indirectly supervised (OR=1.75 [95%CI 1.08 - 2.85] or independent NPC care (OR=2.14 [95%CI 1.05 - 4.34]).

## DISCUSSION

This study of an NPC EC training program in rural Uganda demonstrates that direct supervision by EM physicians of NPC EC can reduce overall mortality. Furthermore, this mortality impact is restricted to the most severely ill subset of patients, with independent NPC care having similar outcomes to care with direct EM physician supervision for the vast majority of patients. These findings are consistent with a prior study by our author group showing the mortality benefit for direct EM physician supervision was restricted to the most severely ill subset of children under 5 years of age.[42] We are not aware of any other studies addressing mortality rates of patients cared for by EC specialty trained NPC in similar LMIC settings. This finding has potentially profound implications for policy to maximize workforce potential in the rapidly developing field of EC in Uganda and in similar settings.

One of the fundamental challenges of our analysis of this NPC training program was the rapidly changing background of the health system in Uganda during the study period (2009-2019). Many of the most profound shifts seen in our study likely reflect the overall changes in Ugandan health care. As shown in Figure 2, overall mortality significantly (p<0.001) decreased by almost 70% during the study period. While impressive, this finding is consistent with the 63% reduction in national crude death rate during the study period.[49] Similarly, we saw many demographic shifts in our population over time (Table 1) including fewer emergencies in children under 5, older patients and reduced rates of malaria. Again, these are consistent with Ugandan national trends over that time period.[49] Figure 2 showed how the patient population was more likely to have normal vital signs and less likely to have abnormal vital signs, therefore being less likely to have severe illness at the time of presentation to the emergency unit. The proportion of patients receiving complete vitals increased over time, so this effect is unlikely due to changing data collection but rather by population level trends in overall health and/or earlier EC health care seeking behavior.

With Ugandans becoming healthier and living longer throughout the decade-long study period, mortality analysis required comparing outcomes for similar patients during different eras of NPC supervision. Our initial analysis focused on grouping patients by their number of abnormal vital signs to compare mortality across program eras (Figure 3). While this was an appealing approach given its simplicity, the findings were conflicting with direct supervision being associated with significantly *decreased* mortality in the highest mortality group but also significantly *increased* mortality in the lowest mortality group.

We therefore developed a logistic regression model for emergency unit mortality to identify the independent contribution of supervision. Within this model, indirect supervision of NPC care and independent NPC care were both seen to have increased mortality when compared to direct supervision of NPC care (“Indirect” OR=1.49 [95%CI 1.07 - 2.09], “Independent” OR=1.76 [95%CI 1.09 - 2.86]). This is an expected finding, as no argument exists in this manuscript or elsewhere suggesting complete equality between physician and NPC training, practice or outcomes. Rather, this finding clearly highlights the importance of the scaling-up of the ongoing EM physician training efforts in Uganda to reduce mortality in EC. While EM physician care for all emergency patients is ideal, the current rate of EM specialist training, health system funding, and high demand for EM specialist physicians at training institutions and in administrative roles, means that the ideal of EM specialist clinical care in emergency units throughout Uganda may be decades away from being realized. Therefore, optimizing the role of NPCs can help address the current gap between EC patients and providers.

Because our vital sign analysis suggested that the benefit of direct supervision was restricted to the highest-risk subset of patients, and this was consistent with prior studies showing the benefit of direct supervision was limited to severely ill pediatric patients, we performed additional sensitivity analysis stratifying by vital signs.[42] We found patients with two or fewer abnormal vital signs had no significant reduction in mortality with direct supervision.

Importantly, there was no trend towards harm from direct supervision as suggested by the crude vital sign analysis. Patients with three or more vital sign abnormalities did have a clinically and statistically significant mortality benefit with direct EM physician supervision. We believe this finding could be used at triage to immediately identify patients most likely to receive benefit from direct EM physician clinical care or direct supervision of NPC care.

Indirect physician supervision did not clearly impact mortality in either crude vital sign or logistic regression analysis. This may stem from several limitations of indirect supervision in the GEC model: GEC only had a volunteer EM physician on site for approximately 85% of the weeks of the indirect era, indirect supervising EM physicians did not have Ugandan medical licenses and were not permitted to provide direct care, and volunteers had with differing levels of training and local expertise. Further, no standardized protocol existed to define patients for which NPCs should involve the EM physician in care. Further study is required to determine if protocolized and consistent indirect physician supervision could provide a mortality reduction for high-risk patients similar to direct physician supervision.

In total, this manuscript shows that direct supervision of NPC care by an EM physician reduces overall mortality. We strongly support the ongoing development of EM specialty training for physicians in Uganda to help achieve the penultimate goal of providing EM physician clinical care or direct supervision for all patients. However, current EC staffing shortages in Uganda and elsewhere is SSA are likely to persist for decades to come and augmenting the physician workforce with EC specialty-trained NPCs — who can be trained more rapidly and at lower cost [19,38,50] — is a clear path forward to addressing the immediate EC needs faced by millions of Ugandans today. Additionally, multiple prior studies have shown the NPCs are much more likely to work in rural areas — where healthcare providers are desperately needed in SSA [4] — when compared to physicians.[19] Our analysis shows that a synergy between these groups is possible: NPCs are capable of safely delivering independent care for less severely ill patients (approximately 90% of patients in our study population) with mortality outcomes similar to care supervised directly by EM physicians, but direct EM physician supervision of NPC care of the most severely ill patients can reduce mortality.

### Limitations

This is a single center, retrospective study of an emergency unit database. Mortality follow-up was limited to three days. While one week and one month mortality is undoubtedly important, three-day follow-up was chosen both to minimize loss to follow-up in a setting where most patients do not have consistent ability to receive phone calls and because follow-up after three days was thought to be less reflective of outcomes related to acute care provided in the emergency unit. Inpatient mortality was affected not just by emergency unit care but also by hospital ward care. However, this care was provided similarly throughout the study, making it unlikely to bias outcomes in comparisons between eras. The decision to code missing blood pressures in children as normal likely biased the study towards seeing pediatric patients as lower risk. However, since those patients were predominantly clustered in the direct supervision era, that decision would have biased results towards the null hypothesis (no impact from direct supervision) and thus was unlikely to bias our findings overall. Lastly, there was a high loss to follow-up in discharged patients over the duration of the study. However, with a mortality rate of 0.08% (n=7 deaths in 9,175 discharges) in those with complete follow-up, it is highly unlikely that the 8,308 discharged patients lost to follow-up represent a significant number of fatal cases excluded from our analysis. The 6.3% loss to follow-up rate for admitted and direct to theatre patients was otherwise considered more than adequate given the challenges of emergency unit data collection in Sub-Saharan Africa.

## CONCLUSIONS

This manuscript shows that task-sharing to address EC staffing shortage with EC specialty-trained NPCs is an efficient and safe way forward. As Uganda strives to reach the goal of consistent EM physician coverage of emergency units, operationalizing a hybrid model with

EM physician supervision of otherwise independent NPC care for the sickest EC patients has the potential to save lives. Based on the robust evidence base we report above, the authors’ recommendations are as follows:

1. Scale up EM physician development and training: The highest risk approximately 10% of patients had nearly a 50% reduction in mortality with physician involvement, and direct supervision significantly reduced overall mortality.
2. Increase capacity for EC NCP training: EC NPCs provided independent care comparable to care given with direct EM physician supervision for approximately 90% of patients over the study period.
3. Create triage protocols for early identification of the highest risk patients: in our analysis patients with hypoxia or with 3 or more abnormal vital signs are at highest risk for mortality and most likely to derive benefit from EM physician clinical care or direct EM physician supervision of NPC care.
4. Create clear protocols and systems to provide EC NPCs with direct supervision in person or via phone/telehealth consultation by EM physician for critically ill and high-risk patients.

## Data Availability

Data sharing is not included in the studies IRB approval

## ACKNOWLEDGMENTS

The authors sincerely thank the Global Emergency Care Collaborative Investigators Mark Bisanzo, Heather Hammerstedt, Stacey Chamberlain and Bradley Dreifuss for their work in developing the EC and research infrastructure that made this study possible. Additionally, we would like to thank the GEC Research Team Members Charles Ndyamwijuka, Adrine Kusasira and Nelly Mbabazi for their tireless work on the research tool, interviews and data analysis, Hilary Kizza, GEC Program Coordinator, for his administrative support and Edgar Mugema Mulogo for his assistance conceptualizing this project in the Ugandan context. The Hospital Management Team at Karoli Lwanga Hospital also provided invaluable support for this study and the ECP program. Finally, we would like to thank the Emergency Care Practitioners for their assistance with patient recruitment and data analysis, as well the critical EC they provided.

